# Differential occupational risks to healthcare workers from SARS-CoV-2: A prospective observational study

**DOI:** 10.1101/2020.06.24.20135038

**Authors:** David W Eyre, Sheila F Lumley, Denise O’Donnell, Mark Campbell, Elizabeth Sims, Elaine Lawson, Fiona Warren, Tim James, Stuart Cox, Alison Howarth, George Doherty, Stephanie B Hatch, James Kavanagh, Kevin K Chau, Philip W Fowler, Jeremy Swann, Denis Volk, Fan Yang-Turner, Nicole E Stoesser, Philippa C Matthews, Maria Dudareva, Timothy Davies, Robert H Shaw, Leon Peto, Louise O Downs, Alexander Vogt, Ali Amini, Bernadette C Young, Philip Drennan, Alexander J Mentzer, Donal Skelly, Fredrik Karpe, Matt J Neville, Monique Andersson, Andrew J Brent, Nicola Jones, Lucas Martins Ferreira, Thomas Christott, Brian D Marsden, Sarah Hoosdally, Richard Cornall, Derrick W Crook, David Stuart, Gavin Screaton, Oxford University Hospitals Staff Testing Group, Timothy EA Peto, Bruno Holthof, Anne-Marie O’Donnell, Daniel Ebner, Christopher P Conlon, Katie Jeffery, Timothy M Walker

**Author notes:** **Corresponding author:** Dr David Eyre. Contributed equally (Katie Jeffery, Timothy M Walker).

## Abstract

**Background:** Personal protective equipment (PPE) and social distancing are designed to mitigate risk of occupational SARS-CoV-2 infection in hospitals. Why healthcare workers nevertheless remain at increased risk is uncertain.

**Methods:** We conducted voluntary Covid-19 testing programmes for symptomatic and asymptomatic staff at a UK teaching hospital using nasopharyngeal PCR testing and immunoassays for IgG antibodies. A positive result by either modality determined a composite outcome. Risk-factors for Covid-19 were investigated using multivariable logistic regression.

**Results:** 1083/9809(11.0%) staff had evidence of Covid-19 at some time and provided data on potential risk-factors. Staff with a confirmed household contact were at greatest risk (adjusted odds ratio [aOR] 4.63 [95%CI 3.30-6.50]). Higher rates of Covid-19 were seen in staff working in Covid-19-facing areas (21.2% vs. 8.2% elsewhere) (aOR 2.49 [2.00-3.12]). Controlling for Covid-19-facing status, risks were heterogenous across the hospital, with higher rates in acute medicine (1.50 [1.05-2.15]) and sporadic outbreaks in areas with few or no Covid-19 patients. Covid-19 intensive care unit (ICU) staff were relatively protected (0.46 [0.29-0.72]). Positive results were more likely in Black (1.61 [1.20-2.16]) and Asian (1.58 [1.34-1.86]) staff, independent of role or working location, and in porters and cleaners (1.93 [1.25-2.97]). Contact tracing around asymptomatic staff did not lead to enhanced case identification. 24% of staff/patients remained PCR-positive at ≥6 weeks post-diagnosis.

**Conclusions:** Increased Covid-19 risk was seen in acute medicine, among Black and Asian staff, and porters and cleaners. A bundle of PPE-related interventions protected staff in ICU.

## Introduction

On 23^rd^ March 2020 the UK followed other European countries in locking down its population to mitigate the impact of the rapidly evolving Covid-19 pandemic. By 5^th^ May the UK had recorded Europe’s highest attributed death toll.(1)

Lock-down isolated many UK households but staff maintaining healthcare services continued to be exposed to patients and other healthcare workers (HCW). National Health Service (NHS) hospitals endeavoured to provide personal protective equipment (PPE) in line with Public Health England (PHE) guidelines in clinical areas and encouraged social distancing elsewhere. Despite these measures the incidence of Covid-19 among HCWs is higher than in the general population.(2)(3)

Multiple studies have investigated Covid-19 in HCWs.(2,4–6) However, crucial to designing a safe working environment and maintaining effective healthcare services is an understanding of the risks associated with specific roles and to individuals, and whether risk is associated with social-mixing, direct exposure to Covid-19 patients or PPE type. Some studies have suggested exposure to Covid-19 patients poses increased risk,(2,7,8) whilst others have not.(9–11) However, none have addressed these questions by comprehensively investigating all staff groups across an institution, simultaneously assessing symptomatic and asymptomatic incidence.

Oxford University Hospitals NHS Foundation Trust (OUH) has offered SARS-CoV-2 PCR and antibody testing to all staff to improve infection prevention and control for staff and patients. We present the results of this large, high-uptake programme.

## Methods

### Setting and data collection

OUH spans four teaching hospitals with 1000 beds and 13,800 staff, serving a population of 680,000 and acting as a regional referral centre. The first patients with Covid-19 were admitted to OUH in mid-March 2020. SARS-CoV-2 testing, initially reserved for in-patients, was extended to symptomatic staff and household contacts with fever or new-onset cough from 27^th^ March. These PCR results are presented.

A voluntary asymptomatic screening programme for all staff working anywhere on site commenced on 23^rd^ April. Naso- and oro-pharyngeal swabs were obtained for real-time-PCR for SARS-CoV-2 and blood for serological analysis. Data on potential Covid-19 risk factors were collected (see Supplement). Results are presented for the first asymptomatic sample from each staff member.

### Infection control

From 1^st^ February 2020, “level-2 PPE” (FFP3/N99 mask, eye protection, gown, gloves) was mandated for any contact with a confirmed or suspected case. From 8^th^ March this was downgraded to “level-1 PPE” (surgical mask, optional eye protection, apron, gloves), except for aerosol generating procedures.(12) From 1^st^ April a minimum of level-1 PPE was mandated for all patient care, regardless of Covid-19 status (Supplementary Table S1).

### Laboratory assays

SARS-CoV-2 PCR was performed using a PHE-designed RdRp assay and several commercial platforms. Serology for SARS-CoV-2 IgG to nucleocapsid and trimeric spike were performed using the Abbott Architect i2000 chemiluminescent microparticle immunoassay (CMIA) and an enzyme-linked immunosorbent assay (ELISA) developed at the University of Oxford (Supplement).(13)

### Ethics

All asymptomatic staff data collection and testing were part of enhanced hospital infection prevention and control measures instituted by the UK Department of Health and Social Care. Deidentified data from staff testing and patients were obtained from the Infections in Oxfordshire Research Database (IORD) which has generic Research Ethics Committee, Health Research Authority and Confidentiality Advisory Group approvals (19/SC/0403, ECC5-017(A)/2009). De-identified patient data extracted included admission and discharge dates, ward location and positive Covid-19 test results.

### Statistical analysis

Univariable and multivariable logistic regression were performed to assess risk factors for infection and to determine associations between self-reported symptoms and Covid-19. A composite end point of ‘Covid-19 at any time’ of a positive PCR test or detection of IgG by ELISA and/or CMIA was used. Univariable and multivariable linear regression were used to assess the relationship between ward-based Covid-19 patient infectious pressure and the proportion of infected staff working on that ward (Supplement).

## Results

By mid-March 2020 OUH saw daily admissions of patients with Covid-19. By 8^th^ June, 636 patients had been admitted within a week of a confirmed Covid-19 diagnosis. Incidence peaked during the week beginning 30^th^ March (n=136/week, Figure 1). Routine testing of symptomatic staff began on 27^th^ March; incidence peaked the week beginning 6^th^ April (n=98/week). By 8^th^ June 348/1498 (23%) symptomatic staff tested were PCR-positive (2.5% of 13,800 OUH staff). Ten staff were admitted with Covid-19 (0.07%); four died (0.03%).

**Figure 1.**
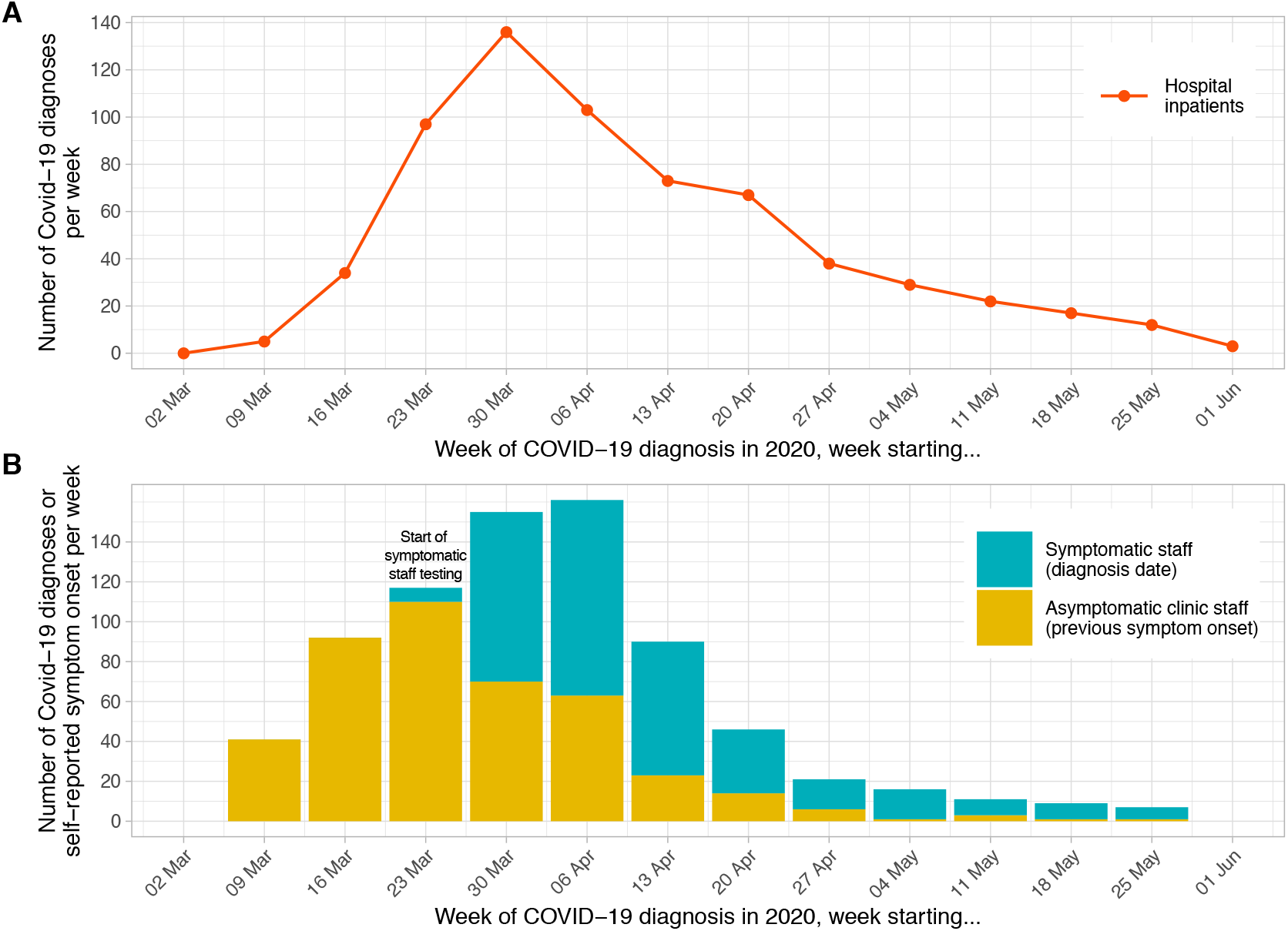
Epidemiological curve – hospital inpatients (panel A) and staff (panel B) diagnosed with Covid-19, by week. Each patient admitted to hospital with a diagnosis of Covid-19 within ±7 days of any day during their admission is plotted based on the date of their positive PCR test. Testing for symptomatic staff was made available from 27^th^ March 2020; staff were asked to attend on day 2-4 of symptoms and are plotted in the week of their positive test. Of 1083 staff positive by PCR or serology at the asymptomatic staff clinic, 183 had been previously diagnosed at the symptomatic staff clinic. Of the remaining 900 positive staff, 472 (52%) reported a date when they believed a Covid-19 illness had begun, these are plotted in yellow above, many with symptoms before the availability of staff testing. As 428 (48%) of staff did not provide a date of symptom onset the true values for the yellow bars on the y-axis are likely to be around 2 times higher.

### Asymptomatic staff testing

Between 23^rd^ April-8th June, 10,610/13,800 (77%) staff registered for asymptomatic testing and 9809(71%) were tested at least once, 9722 by PCR and 9456 by serology. 288/9722 (3.0%) were PCR-positive on their first asymptomatic screen; 130/288 (45%) were assessed to have a new infection and self-isolated. 145 were permitted to remain at work: 61 (21%) tested positive >7 days previously and had recovered and 84 (29%) had a history suggestive of previous Covid-19. Documentation was incomplete for six staff and seven could not be contacted.

### Duration of PCR positivity

Duration of PCR positivity was calculated from staff and patients with consecutive tests. Fewer staff than patients were persistently positive at 7-13 days (exact p=0.003), but results were similar by 14-20 days, 68/159 (43% [95%CI 35-51%]) overall. 34/141 (24% [17-32%]) samples taken after ≥42 days were positive (Figure 2).

**Figure 2.**
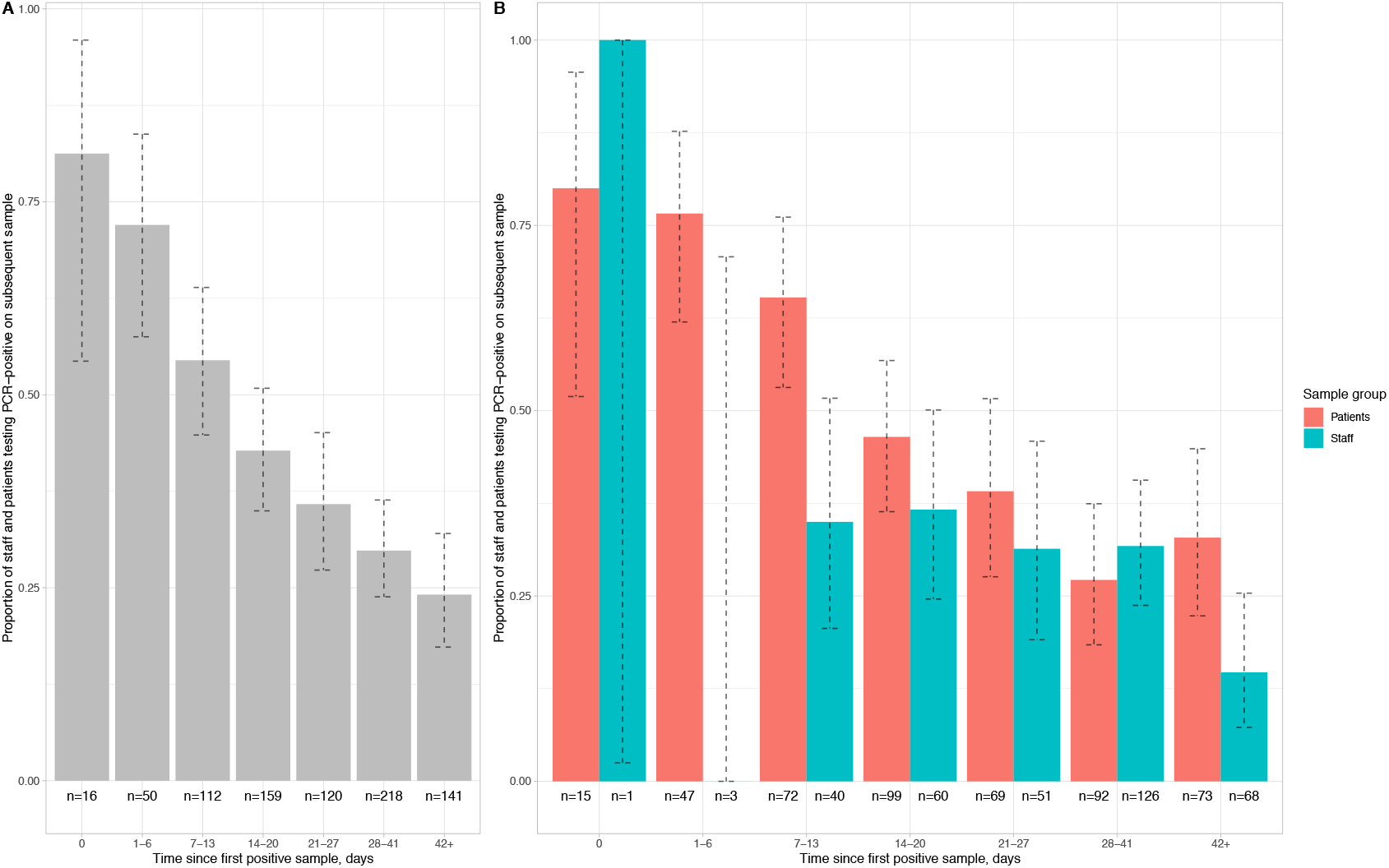
Proportion of staff and patients remaining PCR-positive on repeat nasopharyngeal swabs. Panel A shows pooled data and Panel B data separately for staff and patients. The number of individuals with a repeat test in each time interval is shown below each bar and 95% exact binomial confidence intervals are plotted. All tests following a first positive sample are included up until the first negative sample per patient. The number of tests positive after a repeat swab on the same day is indicative of the sensitivity of a single swab, 15/16 of these swabs were obtained from patients on wards by any available staff member, whereas staff sampling was undertaken by specially trained teams.

### Serology results, symptoms and risk factors

1083/9809 (11.0%) staff attending for asymptomatic screening were positive by PCR or serology, indicating Covid-19 at some time, including 183 previously diagnosed via symptomatic staff testing. 1016/9456 (10.7%) staff with an ELISA/CMIA result were IgG-positive (Table S2).

We asked all staff attending asymptomatic screening about possible Covid-19-related symptoms since 1^st^ February 2020 (Table 1). In a multivariable model containing all symptoms, anosmia or loss of taste was most strongly predictive of Covid-19 (aOR 17.7 [95%CI 14.1-22.3], p<0.001). Other independent predictors included myalgia, fever and cough. Adjusting for other symptoms, sore throat was a negative predictor for Covid-19.

**Table 1.** Association of self-reported symptoms and Covid-19 infection in hospital staff. **\***All interactions with an interaction Wald p values <0.01 are shown.

| Symptom | Symptom reported |  |  |  | Symptom not reported |  |  |  | Univariable |  | Multivariable |  |
| --- | --- | --- | --- | --- | --- | --- | --- | --- | --- | --- | --- | --- |
|  | n | Covid-19 positive | Covid-19 negative | % positive | n | Covid-19 positive | Covid-19 negative | % positive | OR (95% CI) | p value | OR (95% CI) | p value |
| Anosmia or loss of taste | 841 | 477 | 364 | 56.7 | 8966 | 604 | 8362 | 6.7 | 18.1 (15.5-21.3) | <0.001 | 17.7 (14.1-22.3) | <0.001 |
| Myalgia | 1754 | 480 | 1274 | 27.4 | 8053 | 601 | 7452 | 7.5 | 4.7 (4.1-5.3) | <0.001 | 2.0 (1.6-2.4) | <0.001 |
| Fever | 1430 | 391 | 1039 | 27.3 | 8377 | 690 | 7687 | 8.2 | 4.2 (3.6-4.8) | <0.001 | 1.4 (1.2-1.8) | <0.001 |
| Nausea or vomiting | 409 | 129 | 280 | 31.5 | 9398 | 952 | 8446 | 10.1 | 4.1 (3.3-5.1) | <0.001 | 1.3 (1.0-1.7) | 0.08 |
| Cough | 1772 | 395 | 1377 | 22.3 | 8035 | 686 | 7349 | 8.5 | 3.1 (2.7-3.5) | <0.001 | 1.3 (1.1-1.6) | 0.01 |
| Hoarseness | 635 | 132 | 503 | 20.8 | 9172 | 949 | 8223 | 10.3 | 2.3 (1.9-2.8) | <0.001 | 1.2 (0.8-1.7) | 0.36 |
| Shortness of breath | 999 | 235 | 764 | 23.5 | 8808 | 846 | 7962 | 9.6 | 2.9 (2.5-3.4) | <0.001 | 1.2 (0.9-1.5) | 0.34 |
| Diarrhoea | 595 | 143 | 452 | 24.0 | 9212 | 938 | 8274 | 10.2 | 2.8 (2.3-3.4) | <0.001 | 1.2 (0.9-1.5) | 0.31 |
| **Fatigue** | 2660 | 575 | 2085 | 21.6 | 7147 | 506 | 6641 | 7.1 | 3.6 (3.2-4.1) | <0.001 | 1.0 (0.8-1.3) | 0.96 |
| **Nasal congestion** | 1828 | 347 | 1481 | 19.0 | 7979 | 734 | 7245 | 9.2 | 2.3 (2.0-2.7) | <0.001 | 1.0 (0.8-1.2) | 0.89 |
| **Sore throat** | 2200 | 349 | 1851 | 15.9 | 7607 | 732 | 6875 | 9.6 | 1.8 (1.5-2.0) | <0.001 | 0.7 (0.5-0.8) | <0.001 |
| **\*Hoarseness + Anosmia**<br>**or loss of taste** |  |  |  |  |  |  |  |  |  |  | 0.5 (0.3-0.8) | 0.003 |
| **\*Shortness of breath +**<br>**Anosmia or loss of taste** |  |  |  |  |  |  |  |  |  |  | 0.4 (0.3-0.7) | <0.001 |

Prior to asymptomatic testing, 537/1081 (49.7%) staff subsequently testing positive thought they had already had Covid-19, compared to 1092/8726 (12.5%) testing negative. 64/170 (37.6%) staff reporting household contact with a PCR-confirmed case tested positive, compared to 1017/9637 (10.6%) without (p<0.001). SARS-CoV-2 infected staff were also more likely to report suspected, but unconfirmed contacts, and non-household contacts (Figure 3, Table S3). 354/2119 (16.7%) staff reporting workplace contact without PPE with a known or suspected Covid-19 patient tested positive, compared with 727/7688 (9.5%) not reporting similar exposure. To mitigate recall bias we repeated this analysis restricted to staff who did not think they had had Covid-19: 56/1618 (9.6%) reporting an exposure were positive compared to 388/6560 (5.9%) who did not (p<0.001).

**Figure 3.**
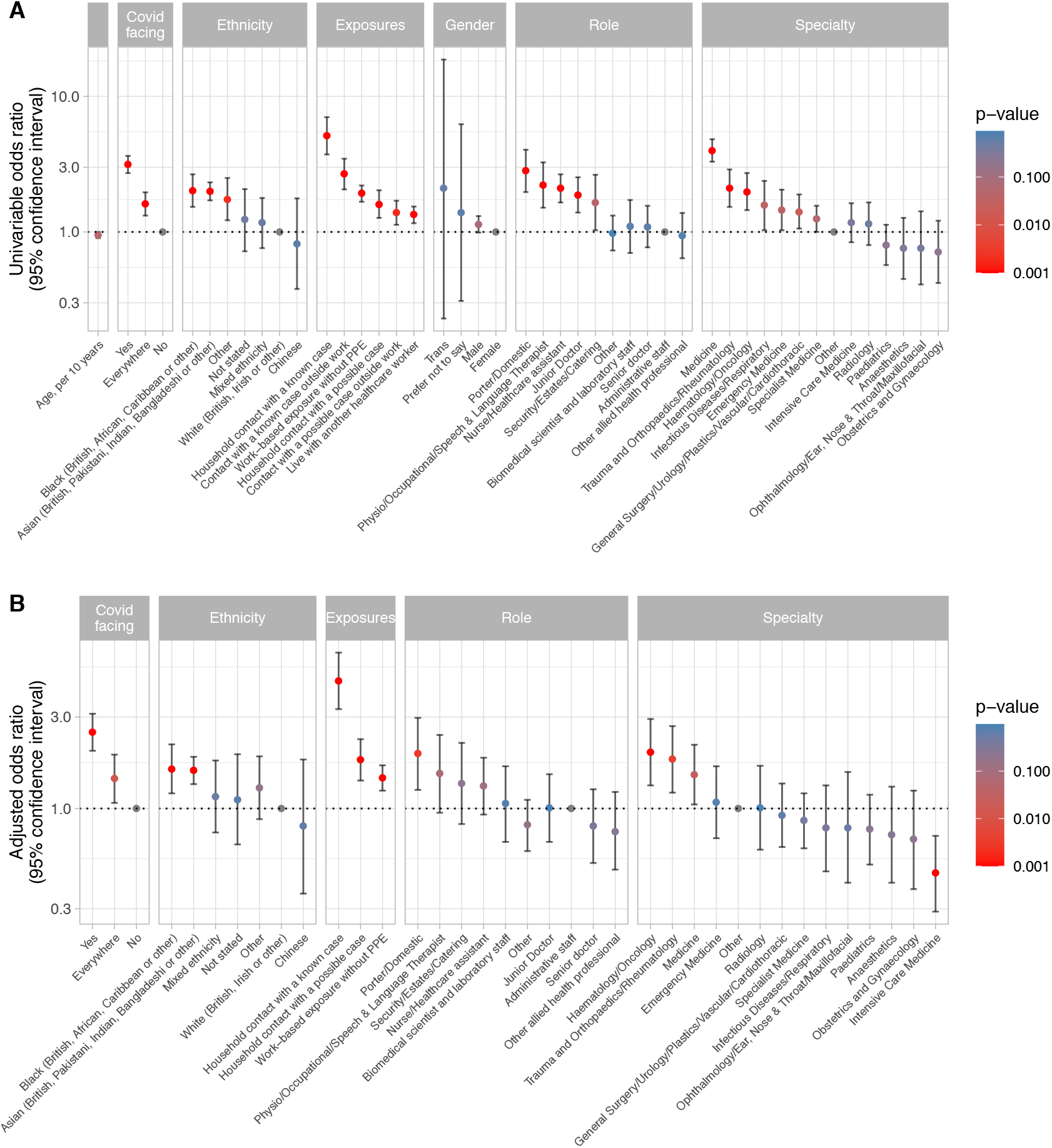
Univariable (panel A) and multivariable (panel B) relationships between risk factors and staff infection with SARS-CoV-2. See Supplementary Table S3 for count data, univariable and multivariable odds ratios. Pairwise interactions were sought between all variables the multivariable model, a single interaction exceeded the p<0.01 screening threshold, representing decreased risk of Covid-19 in emergency department staff reporting exposure to a Covid-19 without PPE (p=0.002). However, given the large number of interactions sought and biological implausibility the interaction is omitted from the model presented. For the purpose of plotting p values <0.001 were rounded up to 0.001. No data were available for 2 staff members.

We further investigated risk of workplace Covid-19 acquisition. 362/1718 (21.1%) staff on wards caring for patients with Covid-19 were infected, compared to 585/7028 (8.2%) on non-Covid-19 facing wards/other areas, and 136/1063 (12.8%) staff working across multiple areas (p<0.001). Covid-19 facing areas included the emergency department, acute medical and surgical wards, the respiratory high dependency unit (HDU) and three intensive care units (ICUs). However, the proportion of staff with a positive test working in acute medicine (212/775, 27.4%) was greater than in the emergency department (41/340, 12.1%) and in the ICUs (43/434, 9.9%) (Figure 3A, S1, Table S3).

Rates of Covid-19 infection varied by staff role: porters and cleaners had the highest rates (58/323, 18.0%), followed by physio-, occupational and speech and language therapists (45/307, 14.7%) and nurses/healthcare-assistants (543/3886, 14.0%). Junior medical staff had higher rates (104/820, 12.7%) than senior medical staff (54/692, 7.8%). Administrative staff had the lowest proportion (86/1196, 7.2%) of any major staff group (Figure 3A, S2, Table S3).

There was limited evidence that male staff were more at risk of infection than female staff (301/2506 [12.0%] positive vs. 779/7284 [10.7%], p=0.07) and risk decreased with age (univariable odds ratio [OR], per 10 years, 0.95 [95%CI 0.90-1.00, p=0.05], Figure S4). Covid-19 rates varied by self-described ethnicity. 654/7064 (9.3%) staff describing themselves as White (British/Irish/other) were infected, compared to 279/1649 (16.9%) and 65/381 (17.1%) staff describing themselves as Asian (British/Pakistani/Indian/Bangladeshi/other) or Black (British/African/Caribbean/other) respectively. Rates in staff describing themselves of mixed ethnicity or Chinese were 25/235 (10.6%) and 7/91 (7.7%) (Figure 3A, S3, Table S3).

### Multivariable analysis

In multivariable analysis (Figure 3B, Table S3), controlling for factors including hospital-based Covid-19 exposure, role, specialty and ethnicity, household contact with known (adjusted OR [aOR] 4.63, 95%CI 3.30-6.50, p<0.001) or suspected (1.79, 1.40-2.30, p<0.001) cases remained important risk factors. Working in Covid-19 facing areas (2.49, 2.00-3.12, p<0.001) or throughout the hospital (1.43, 1.07-1.91, p=0.02) was associated with increased risk compared to non-Covid-19 areas, as was workplace-based exposure to a suspected or known Covid-19-positive patient without PPE (1.44, 1.24-1.68, p<0.001). The latter could not be entirely accounted for by recall-bias as the association persisted restricting to staff who did not think they had had Covid-19 (1.28, 1.04-1.57, p=0.02).

Risk of Covid-19 infection varied by speciality, even after accounting for working in a Covid-19 facing area. Those working in acute medicine were at increased risk, (aOR 1.50, 95%CI 1.05-2.15, p=0.03), while ICUs were at lower risk (0.46, 0.29-0.72, p=0.001). Increased risk was also seen in in orthopaedics and haematology, reflecting staff-based outbreaks as these wards saw very few Covid-19 patients. The greatest risk of infection by role remained for porters and cleaners (1.93, 1.25-2.97, p=0.003). By ethnic group, Black (1.61, 1.20-2.16, p=0.001) and Asian (1.58, 1.34-1.86, p<0.001) staff were at greatest risk of Covid-19.

### Heterogeneity between hospitals and wards

We investigated the relationship between infectious pressure from patients and the proportion of staff infected by considering each admitted patient infectious from -2 to +7 days around their first positive SARS-CoV-2 PCR. At a hospital building level (Figure 4A), the two buildings admitting most patients with Covid-19 had higher levels of staff infection (13.8%, 15.0%) than the majority of other buildings (5.1-8.5%). However, one site with low rates of patient infection and another, non-clinical site without patients had rates of 13.0% and 20.3% respectively. At a ward level (Figure 4B), there was only a weak positive correlation between Covid-19 pressure from patients and staff infection rates (R^2^=0.07, p=0.02). ICUs and the HDU had lower rates of staff infection for a given Covid-19 pressure than general Covid-19 facing wards (linear regression coefficient -28% [95%CI -45%, -11%; p=0.002]). While dedicated Covid-19 cohort wards had similar rates of staff Covid-19 to general wards overall (Table S4), several general wards had much higher rates (Figure 4B).

**Figure 4.**
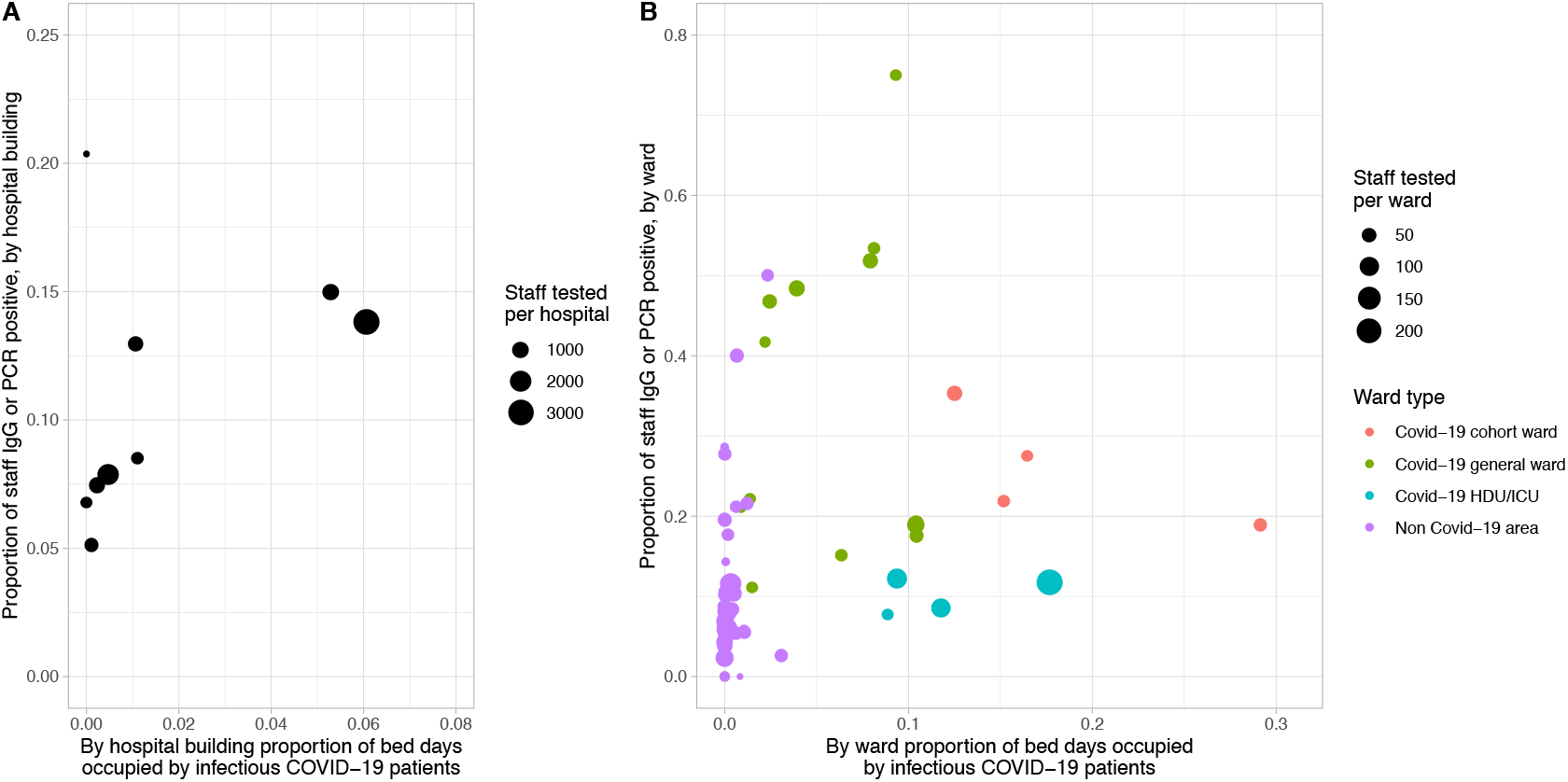
Proportion of staff infected by extent of Covid-19 infectious pressure from patients, by 8 hospital buildings across 4 hospitals (panel A) and by ward (panel B). Covid-19 infectious pressure was calculated by considering each patient infectious from -2 to +7 days around the date of their first positive SARS-CoV-2 PCR test. Only staff working in a single hospital or ward are included in the plot. Wards with fewer than 10 staff tested are not plotted. Covid-19 cohort wards admitted only patients with suspected or known Covid-19, whereas Covid-19 general wards were acute medical wards receiving new admissions and acute medical patients initially believed not to have Covid-19. Non Covid-19 areas did not admit suspected Covid-19 patients and any suspected or confirmed Covid-19 patients were transferred off these wards as soon as possible.

### Contact tracing

PCR-positive asymptomatic staff who had not previously had Covid-19 were asked to name all colleagues with whom they had had >5 minutes of face-to-face conversation or been within 2 meters for >15 minutes, within the past 48 hours, without a face mask. During the first two weeks of asymptomatic screening, 130 contacts were tested 7 days after contact with their index case, and 62 re-attended at day 14. Only one contact tested positive. As this rate of detection was below the background rate, contact tracing was discontinued for asymptomatic staff.

## Discussion

We present the results of a large and comprehensive Covid-19 staff testing programme across four teaching hospital sites in one UK county, attended by 71% of 13,800 staff. Using a composite outcome of either a positive PCR or serology result, by June 8^th^ we detected evidence of Covid-19 at some time in 11% of staff. Put in context, UK-wide seroprevalence was 6.8% on 28^th^ May 2020, with a higher incidence among healthcare workers than in the general population.(14)

We observed varying risk to our hospital staff associated with working location, occupational role and demographic factors. The greatest risk was associated with Covid-19 infected household contacts (although only 36.7% of staff with a contact became infected) and with working in Covid-19-facing areas (21.2% vs. 8.2% elsewhere) where there was 1 additional SARS-CoV-2 infection per ∼8 staff compared to elsewhere. On univariable analysis staff with most direct patient contact were at increased risk including porters, cleaners, nurses, healthcare-assistants, therapists and junior doctors. Adjusting for working in a Covid-19 area captured much of this risk, except for porters and cleaners who had the highest adjusted risk of any staff group, and who typically operate across the hospital.

A heterogenous pattern also emerged across different Covid-19-facing areas. Risk seen on acute medical wards was greater than in the emergency department which was often bypassed by Covid-19 patients, whilst working on a Covid-19 facing ICU was relatively protective. One key difference across these areas was the type of PPE worn and the time periods over which it was mandated. Level-2 PPE was mandatory on ICU and HDU throughout whereas policies changed over time on other wards (Table S1). Moreover, staff on ICU and HDU received extensive training in donning and doffing and had dedicated space and supervision for this whereas ward staff did not. Prior to 1^st^ April 2020, in line with national guidance, in acute medical areas outside of Covid-19 cohort wards level-1 PPE was only worn for contact with patients with known or suspected Covid-19, potentially leading to unprotected exposure to patients in whom Covid-19 was not suspected, such as afebrile elderly patients with delirium, functional decline or diarrhoea. This likely explains the greater number of staff infected in several acute medical wards (shown in green near the top of Figure 4B), compared to Covid-19 cohort wards (shown in red).

The reported rates of exposure without PPE were similar among medical and ICU staff (42% and 38%, Table S5), likely reflecting exposures to ICU staff visiting wards to assess critically ill patients. Universal admission testing was only introduced on 24th April 2020, and the limited availability and speed of testing in the early phase of the pandemic likely delayed identification of some Covid-19 cases.

It is difficult to say whether level-1 PPE was less protective than level-2. Increased Covid-19 in staff reporting exposure to a Covid-19 patient without PPE suggests surgical masks afford some protection, and protection from influenza has been reported to be similar using surgical versus FFP2 masks.(15) However, it is likely that a bundle of measures (level-2 PPE, training, supervision and space for donning and doffing, increased staffing levels) influenced the lower risk in ICU and HDU staff (Figures 3, 4). As with many infection control intervention-bundles it is difficult to distinguish which component was most important.

It is also likely that staff-to-staff transmission amplified incidence, based on high Covid-19 rates in several wards without large numbers of Covid-19 patients. Future viral genome sequencing studies may allow analysis of the relative contribution of patients and staff to transmission.

Increased risk of adverse outcomes has been widely reported in Black and Asian ethnic groups,(16) with evidence they are also at increased risk of infection.(3,17) Here we show Black and Asian staff were at greater risk of infection after controlling for age, gender, working location, role, and exposure at home. Job role can be thought of as a proxy for socio-economic background but we were not able to control directly for income levels, home circumstances, pre-morbid conditions or other potential structural inequalities. That staff working as porters or cleaners had the greatest adjusted risk of infection is consistent with economics playing a part in risk, potentially reflecting conditions outside of the hospital, e.g. dense occupancy of living space due to lower incomes.

To calculate a personalised Covid-19 risk score, all factors in the multivariable model need to be considered, e.g. an Asian Covid-19-facing medical nurse is 7.74 (95%CI 5.65-10.61) more likely to be infected than a white non-Covid-19-facing administrative worker. Notably, this exceeds the risk of living with someone with known Covid-19 (aOR 4.63, 95%CI 3.30-6.50).

Limitations of our study include its cross-sectional nature and that data gathered on particular exposures may be subject to recall bias. It is unknown what proportion of staff were infected who either mounted no detectable antibody response or in whom it had waned by the time of testing. Our data are also from a single setting and findings may vary by practice, geography and population-wide Covid-19 incidence.(5,6)

Our study suggests that an earlier move to universal level-1 PPE may have prevented some infections, and that a consistent bundle of level-2 PPE provision and use, training, and supervision and space for donning and doffing protected staff working in high-risk areas. Wider deployment of this bundle should be considered where staff are at increased risk. Our study provides data to inform risk assessments for staff, to ensure those staff most at risk are deployed appropriately. Given likely staff-to-staff transmission where COVID-19 patient pressure was low, there is a need to protect all staff regardless of role. This includes reinforcement of measures to support social distancing and raises questions about the role of social inequality in Covid-19 transmission. If some staff are already immune the impact of any future Covid-19 surge may be less marked for staff, although differential deployment or use of PPE based on immune status would require evidence it was safe and socially acceptable. Our testing programme has been highly popular with staff, ensured enhanced detection of those with Covid-19, and now also provides a large cohort to inform studies on the extent of antibody-mediated protection against future infection.

## Data Availability

Data will be made available on request subject via the Infections in Oxfordshire Research Database subject to applicants meeting the ethical and governance requirements of the Database.

## Acknowledgements

We are extremely grateful to all the NHS staff who participated in our programme and provided data and samples. We would like to pay tribute to all the staff working at the Oxford University Hospitals NHS Foundation Trust, and their families, and in particular to those who became seriously unwell and the four staff members who died from Covid-19.

## Declarations

DWE has received lecture fees from Gilead, outside the submitted work. No other author has a conflict of interest to declare.

